# Concordance in pregnancy planning, preconception health behaviours and health information-seeking of pregnant couples: a cross-sectional study

**DOI:** 10.64898/2026.07.01.26356192

**Authors:** Tristan Carter, Danielle Schoenaker, Georgia Marron, Ludivine Colas, Amie Steel

## Abstract

**Introduction:** Relational dynamics between partners within a couple planning pregnancy are critical to their preconception health behaviour change and are largely underexamined. Given the need for both reproductive partners to engage in beneficial preconception health behaviours and the influence couples have on each other’s behaviour, this study examines the concordance between pregnancy planning and preconception health behaviours and health information-seeking within reproductive partner dyads.

**Methods:** A retrospective observational study was undertaken utilizing data from two online cross-sectional 80-item surveys administered simultaneously between December 2020 and September 2021. Eligible study participants were females or birthing people aged 18-49 living in Australia during any stage of pregnancy, and reproductive partners of those that met these criteria. The survey items covered sociodemographic characteristics, level of pregnancy planning, preconception health behaviours, health information seeking, and health history. Cohen’s kappa (K) (categorical variables) and interclass correlation coefficients (ICC) (continuous variables) were used to identify agreement within the couples.

**Results:** Eighty matched dyads of pregnant females and non-pregnant partners were included. Concordance in pregnancy planning was fair (K=0.27) and was primarily seen in couples where both partners reported the pregnancy as planned (42.5%) or ambivalent (18.8%). Couples had very low similarity (ICC:0.22) in weekday alcohol consumption 3 months preconception (pregnant females: 1.2 standard drinks per day (SD1.7); non-pregnant partners: 2.5/day (SD3.5)). Approximately one quarter (26.3%) of couples reported similarities in partners attempting and succeeding in preconception health information-seeking 12 months before pregnancy. There was greater concordance in **not discussing** preconception health topics with GPs, including topics explicitly covered within clinical guidelines.

**Conclusion:** There is notable discordance in couple’s preparation for pregnancy in many behaviours relevant to positive pregnancy outcomes, and in their health service engagement and experience. There is a clear need to provide care to both reproductive partners to ensure the best possible outcome for the future generations.

## Introduction

The physical and mental health of both partners in a couple before pregnancy is critically important to fertility, pregnancy, embryo, birth and offspring outcomes [1, 2]. This period before pregnancy can be weeks, months or years, and is commonly referred to as the preconception period [3, 4]. While the evidence supporting the impact of preconception exposures on these diverse outcomes continues to grow, there are still gaps in translation to real-world health behaviour change in the community [5-7]. Reasons for these gaps include broader societal influences, as well as poor awareness among reproductive-aged couples regarding the importance of pregnancy planning and limitations to health systems and services in delivering preconception care [8, 9]. One of the factors underpinning poor community awareness of preconception health risks is a lack of accurate and current information relevant to the diverse needs of the different groups that would benefit from improving preconception health [5]. These knowledge gaps are not only present in the general public but also among health professionals [9], despite the broad range of professions that currently have, or are argued to potentially have, a valuable role in delivering preconception care [7].

While pregnancy is not always planned, Australian research suggests approximately 75% of pregnant females and 65% of males with pregnant partners have planned pregnancies [10, 11]. The definition of pregnancy planning has evolved in the last decade, with validated instruments measuring pregnancy planning encompassing criteria such as: use of contraceptive methods, appropriateness of the timing of the pregnancy within an individual’s life plans, intention to become pregnant, desire for a baby, discussions about pregnant with the reproductive partner, and engagement with beneficial health behaviours aimed at improving pregnancy, birth and offspring outcomes [5, 12]. Recent research has shed more light on the factors influencing this latter criterion of health behaviour change. Specifically, analysis of drivers of preconception behaviour change among pregnant females has found that their partner’s beliefs about the importance of behaviour change is potentially more influential than the pregnant female’s perceptions of a health professional’s views on the behaviour’s importance [13]. Equally, men’s preconception health behaviours also appear to be influenced more by their perceptions of their partner’s more than a health professional’s beliefs [14].

These relational dynamics are critical and are largely underexamined in the context of preconception health and care. Evidence does, however, indicate there is poor inclusion of content relevant to men in preconception health information and poor engagement with men in preconception health services [15, 16]. Given the need for both reproductive partners to engage in beneficial preconception health behaviours and the influence couples have on each other’s behaviour, this study aims to examine the concordance between pregnancy planning and preconception health behaviours and health information-seeking within reproductive partner dyads.

## Methods

### Study design and setting

A retrospective observational study was undertaken utilizing data from two online cross-sectional surveys administered simultaneously between December 2020 and September 2021. One survey was presented to pregnant females and the other survey to their non-pregnant partners. Ethics approval was obtained through the University of Technology Sydney [UTS] Human Research Ethics Committee [HREC] (ETH23-8329). This study adheres to Strengthen The Reporting of Observational studies in Epidemiology [STROBE] guidelines [17].

### Participant recruitment and survey administration

Eligible study participants were females aged 18-49 living in Australia during any stage of pregnancy, or a reproductive partner of someone that met these criteria. Recruitment occurred using targeted advertisement on social media platforms [Twitter, Facebook, Instagram]. The study explicitly included pregnant people and their reproductive partner. The study acknowledged that within the diversity of family relationships, other reproductive arrangements exist but our study explicitly focused on the two individuals providing genetic material for the study. For this reason, our paper refers to the sex of the birthing person (‘pregnant female’) and their reproductive partner (‘non-pregnant partner’ or ‘expectant partner’).

Data were collected using Qualtrics^TM^ online survey platform. Individuals interested in the social media advertisement accessed a screening instrument which was completed prior to survey commencement to ensure they met the study eligibility criteria as participants and to provide their name and email address. Participants who met the eligibility criteria were sent an email containing the survey link and another survey link to be forwarded via email to their reproductive partner. Survey completion time was approximately 30 minutes and respondents had up to 14 days to complete the survey. An incentive in the way of a random prize draw of $100 was offered for survey completion. All participation was voluntary.

### Survey Instrument

#### Measures

Prior to survey development and testing in Qualtrics^TM^, a preliminary paper-based version was designed and piloted by three individuals. This drafted survey in Qualtrics^TM^ was adapted into two separate surveys and the wording tailored for either (i) the pregnant female or (ii) their expectant partners (See Supplementary Files 1 & 2). Both surveys are similar and consist of 80 items across five domains (1) “About you” (sociodemographic characteristics, general health status and health services use); (2) “Your current pregnancy” (retrospective assessment of the degree and timing of pregnancy planning using the validated Australian version of the LMUP; (3) “Your health behaviours” (changes in preconception health and health behaviours relevant to the Australian (for females) and international (for expectant partners) preconception care guidelines; (4) “Prepregnancy health information and advice” (knowledge on the importance of preconception health, the content of preconception care, and the source(s) of this information); and (5) “Your health history” (health and pregnancy history).

#### Participant demographics

The survey requested information regarding gender, age, highest qualification completed, employment status, relationship status, financial manageability, identification as Aboriginal or Torres Strait Islander, English being spoken as a first language, being born in Australia, and having health care card access. Pregnancy planning status was reported using the London Measure of Unplanned Pregnancy (LMUP) which is a validated instrument consisting of six items that measures the level of intention regarding a pregnancy. There are two different versions of the LMUP used in this study: one developed for pregnant females [18], and a second instrument for non-pregnant partners [19]. Both of these have been developed in a UK population and the instrument for pregnant females has been further validated for Australian participants but not the LMUP for non-pregnant partners.

#### Participant health status and health history

The general health status and presence of long-standing illness, disabilities or infirmity were also items included in the survey. In addition, participants reported actions taken during the 6 months before pregnancy, which included getting tested for sexually transmitted diseases (STI), and 12 months before pregnancy - including dental visits, immunisations assessments or seeing any health professional.

#### Participant health behaviours

Survey responses were provided by participants regarding their physical activity behaviours in the 3 months before pregnancy, as well as smoking habits (ever smoked and if so: smoked 3 months before pregnancy) and alcohol intake patterns (ever consumed and if so: consumed 3 months before pregnancy). The survey also identified health behaviours in the 6 months before pregnancy including contraception use and contraception methods alongside weight loss dieting.

#### Participant preconception health information and advice

Survey items of interest were also reported by participants regarding preconception health information access before becoming pregnant including information access, who did the information relate to, and the source of this information [Books, leaflets, internet, magazines, family or friends, GP, midwife, Pharmacist, Naturopath].

#### Health topics discussed with primary care professionals, family, and friends

Health topics discussed with general practitioners, other healthcare professionals (midwife, obstetrician/gynaecologist, or naturopath), and family or friends were also reported by participants. Topics included: eating a healthy diet, immunisations, smoking, alcohol, physical activity, stopping contraception, folic acid, healthy weight, sexually transmitted infections, multivitamins, omega 3 fatty acids, and general contraception or fertility.

### Data Analysis

Data from each survey were transferred into Stata (version 19.5), cleaned and prepared for merging. Couples were allocated a unique identifier that allowed individuals within a dyad to be matched for analysis. Datasets from the two participant categories were then merged by unique identifier to create one response row per couple, incorporating responses items for each participant from a couple within a row.

The London Measure of Unplanned Pregnancy (LMUP) [18, 20] guided the categorisation of pregnancy planning status to report pregnancies as i) planned, ii) ambivalent, or iii) unplanned. However, the LMUP instrument used for this study did not accurately report all six items. As such, the LMUP is this study is based on the five of six items only.

Responses for several variables (age, employment status, financial manageability, highest educational qualification, and general health status, and contraception use) were combined to limit reporting cell size <5.

Couples’ concordance for each response was described as a proportion of the total couples answering the question. For example, couple concordance was determined for being born in Australia that identified proportions of: (1) pregnant females born in Australia with partners also born in Australia, (2) pregnant females *not* born in Australia with partners also *not* born in Australia, (3) pregnant females born in Australia with partners *not* born in Australia, and (4) pregnant females *not* born in Australia with partners born in Australia. Additional analysis calculated the proportion of all couples with full concordance on being born in Australia, and full concordance in not being born in Australia. Cohen’s kappa with 95% confidence intervals were used to descriptively report the level of agreement within the couple for categorical variables accounting for chance [21]. Weighted Cohen’s kappa was applied to ordinal categorial variables. Kappa agreements were reported as slight (< 0.20), fair (0.21-0.40), moderate (0.41–0.60), substantial (0.61–0.80), and almost perfect agreement (>0.80). Agreement was assessed using percent concordance where small cell sizes and imbalanced response distributions of categorical variables limited the interpretability of Cohen’s kappa. Concordance in pairwise correlations for mean scored items were assessed using interclass correlation coefficients categorised as poor (>0.5), moderate (0.5-0.75), good (0.75-0.9) and excellent agreement (>0.9)[22].

## Results

There were 617 pregnant females and 146 expectant partners who completed the respective surveys sufficient to be included in analysis. A total of 80 pregnant females and partners were matched as couples and included in this study.

### Sociodemographic characteristics

State of residence and relationship status were reported identically for all couples. Most lived in Queensland (n=21, 26.2%), Victoria (n=21, 26.2%) or New South Wales (n=20, 25.0%). More couples were married (n=52, 65.0%) than in a de facto relationship (n=25, 31.2%), and a small number identified as not currently in a relationship, or not living with their partner (n=3, 3.7%). A moderate agreement between couples was identified for age (K=0.58; 95% CI 0.46, 0.70), financial manageability (K=0.57; 95% CI 0.41, 0.73), and health care card access (K=0.45; 95% CI 0.22, 0.68) (see Table 1). Slight concordance was identified for being born in Australia (K=0.14; 95% CI 0.01, 0.21). Most pregnant females who did not identify as Aboriginal or Torres Strait Islander (97.3%) were also with a partner who similarly did not identify.

**Table 1:**
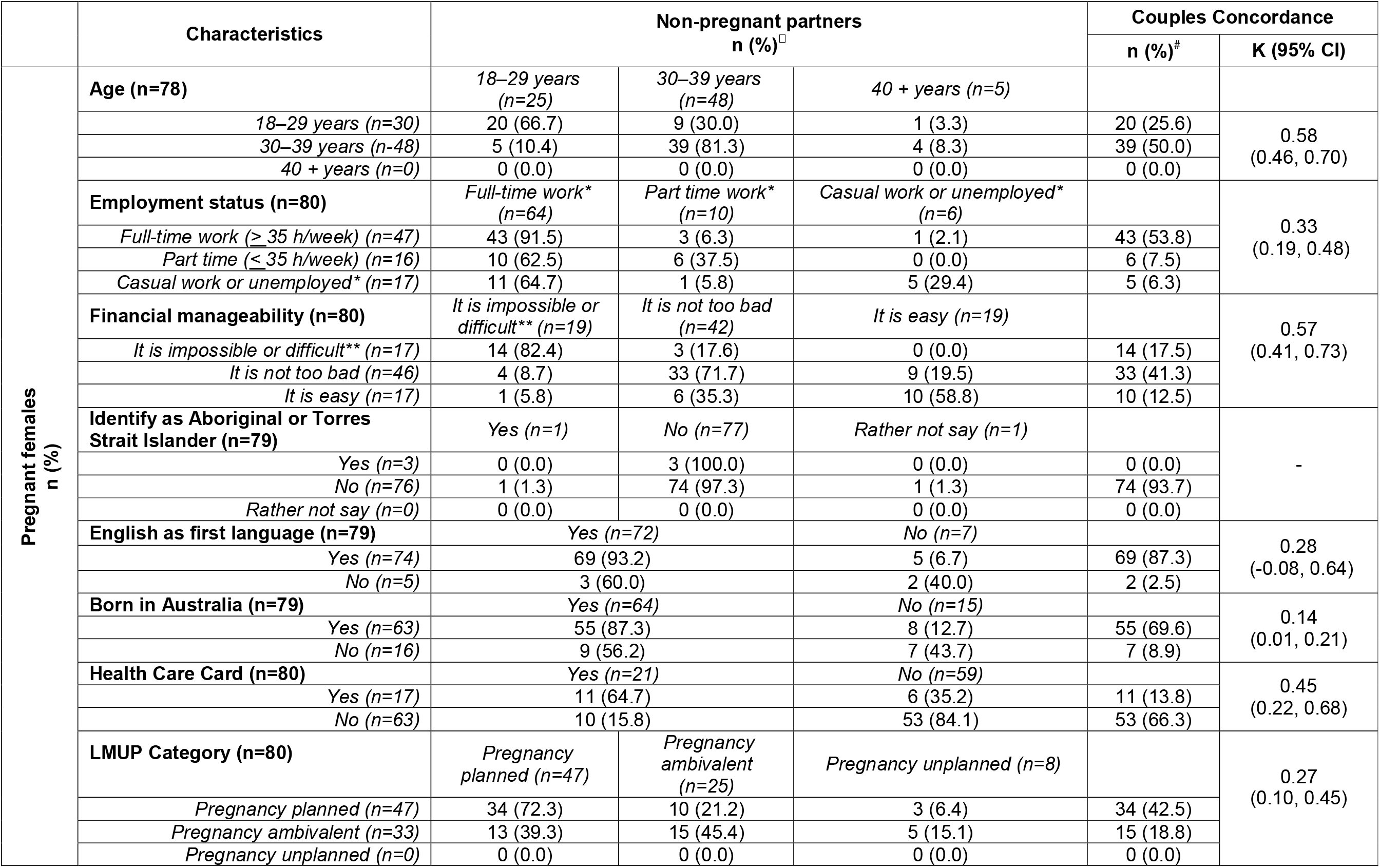
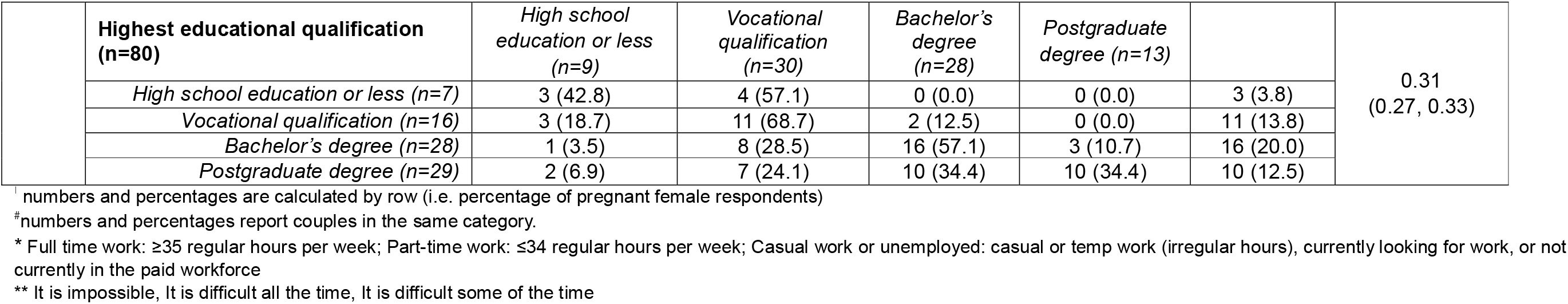
Sociodemographic characteristics of pregnant females and their non-pregnant partners and concordance of characteristics within couples, N = 80.

### Pregnancy planning

Concordance in pregnancy planning was fair (K=0.27; 95% CI 0.10, 0.45), and was primarily seen in couples where both partners reported the pregnancy as planned (42.5%) or ambivalent (18.8%). More than one quarter of pregnant females who indicated a planned pregnancy had a partner who identified as ambivalent (21.2%) or unplanned (6.4%) for the same pregnancy. Among the pregnant females who reported ambivalent pregnancy planning, more than half of non-pregnant partners reported either planned (39.3%) or unplanned (15.1%) pregnancy.

### Participant health status, health history, and health behaviours

Couples had the highest level of concordance for having ever consumed alcohol, demonstrating almost perfect agreement (K=0.82; 95% CI 0.57, 1.00) whereby 90.0% of couples reported both drinking alcohol in the past and 7.1% reported both not ever drinking alcohol (see Table 2). There was moderate agreement between couples on having seen a health professional in the 12 months before becoming pregnant (K=0.52; 95% CI 0.31, 0.73), with more pregnant females consulting a health professional in the 12 months before pregnancy where their partner did not (44.4%), rather than pregnant females not consulting a health professional while their non-pregnant partner did (3.7%). There was fair concordance in having ever smoked (K=0.27; 95% CI 0.04, 0.50) visiting a dentist 6 months before pregnancy (K=0.25; 95% CI 0.04, 0.46) and being tested for a sexually transmitted disease in the same pre-pregnancy time frame (K=0.23; 95% CI 0.02, 0.44). Concordance among smoking or engaging in regular physical activity in the 3 months before pregnancy and among checking immunisation status 12 months before pregnancy did not generate a significant Cohen’s kappa. The percent concordance for smoking indicated all pregnant females who reported smoking had non-pregnant partners reporting the same. In two thirds (66.7%) of couples both partners reported regular physical activity, while 59.5% of couples had both partners report not checking immunisation status.

**Table 2:**
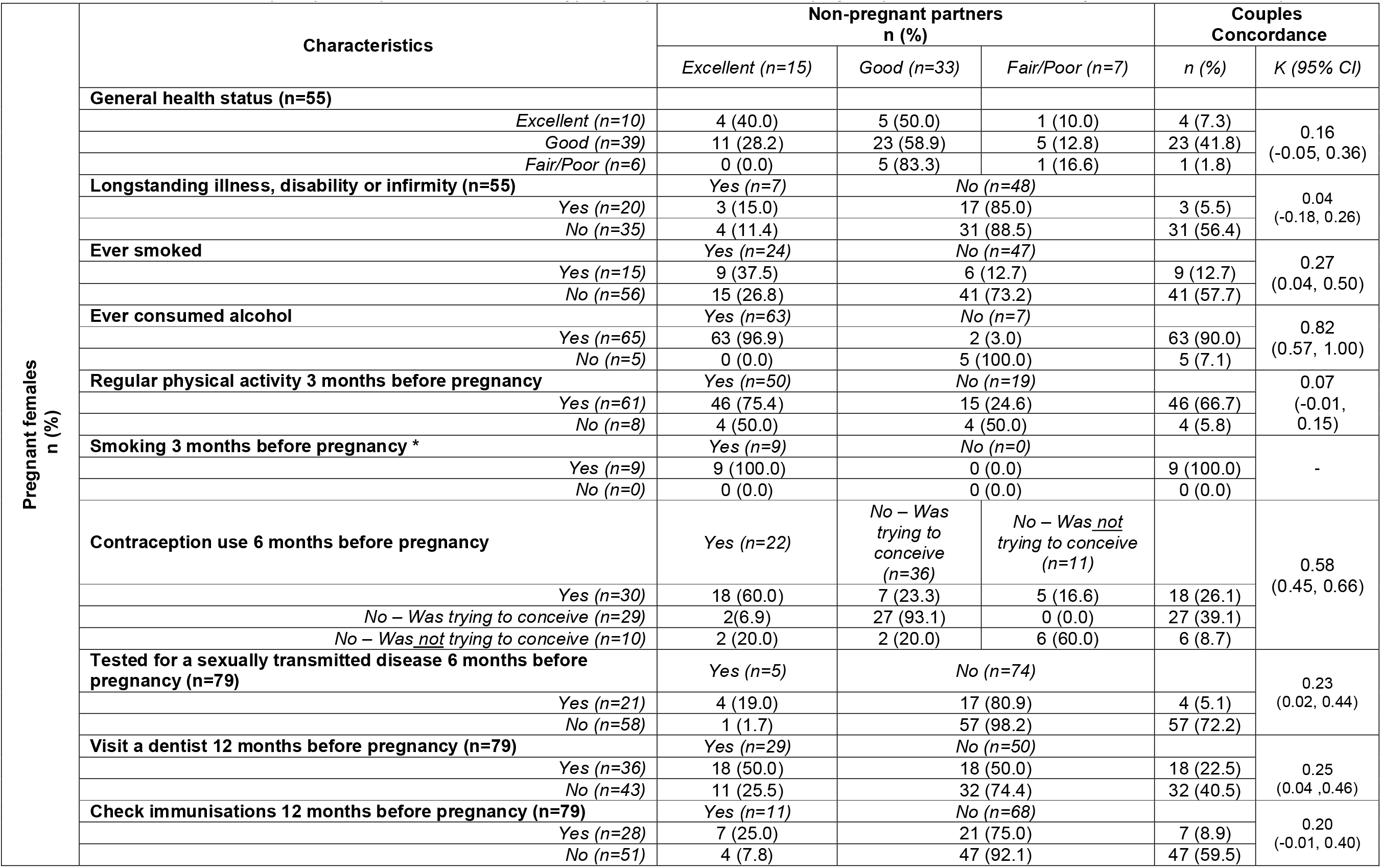
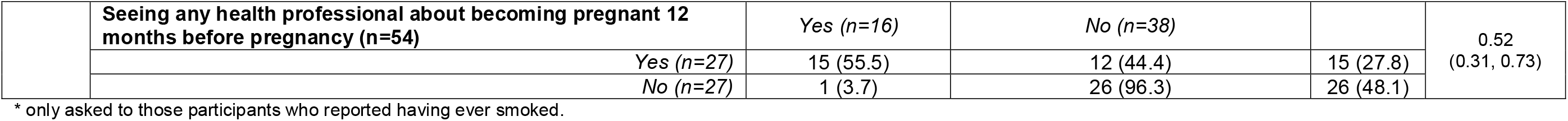
Health status, health history and preconception health behaviours of pregnant females and their non-pregnant partners and concordance of characteristics within couples, N = 80.

|  | Characteristics | Non-pregnant partners<br>n (%) |  |  | Couples<br>Concordance |  |
| --- | --- | --- | --- | --- | --- | --- |
|  |  | Excellent (n=15) | Good (n=33) | Fair/Poor (n=7) | n (%) | K (95% CI) |
| Pregnant females<br>n (%) | **General health status (n=55)** |  |  |  |  |  |
|  | Excellent (n=10) | 4 (40.0) | 5 (50.0) | 1 (10.0) | 4 (7.3) | 0.16<br>(-0.05, 0.36) |
|  | Good (n=39) | 11 (28.2) | 23 (58.9) | 5 (12.8) | 23 (41.8) |  |
|  | Fair/Poor (n=6) | 0 (0.0) | 5 (83.3) | 1 (16.6) | 1 (1.8) |  |
|  | **Longstanding illness, disability or infirmity (n=55)** | Yes (n=7) | No (n=48) |  |  | 0.04<br>(-0.18, 0.26) |
|  | Yes (n=20) | 3 (15.0) | 17 (85.0) |  | 3 (5.5) |  |
|  | No (n=35) | 4 (11.4) | 31 (88.5) |  | 31 (56.4) |  |
|  | **Ever smoked** | Yes (n=24) | No (n=47) |  |  | 0.27<br>(0.04, 0.50) |
|  | Yes (n=15) | 9 (37.5) | 6 (12.7) |  | 9 (12.7) |  |
|  | No (n=56) | 15 (26.8) | 41 (73.2) |  | 41 (57.7) |  |
|  | **Ever consumed alcohol** | Yes (n=63) | No (n=7) |  |  | 0.82<br>(0.57, 1.00) |
|  | Yes (n=65) | 63 (96.9) | 2 (3.0) |  | 63 (90.0) |  |
|  | No (n=5) | 0 (0.0) | 5 (100.0) |  | 5 (7.1) |  |
|  | **Regular physical activity 3 months before pregnancy** | Yes (n=50) | No (n=19) |  |  | 0.07<br>(-0.01, 0.15) |
|  | Yes (n=61) | 46 (75.4) | 15 (24.6) |  | 46 (66.7) |  |
|  | No (n=8) | 4 (50.0) | 4 (50.0) |  | 4 (5.8) |  |
|  | **Smoking 3 months before pregnancy \*** | Yes (n=9) | No (n=0) |  |  | - |
|  | Yes (n=9) | 9 (100.0) | 0 (0.0) |  | 9 (100.0) |  |
|  | No (n=0) | 0 (0.0) | 0 (0.0) |  | 0 (0.0) |  |
|  | **Contraception use 6 months before pregnancy** | Yes (n=22) | No – Was trying to conceive (n=36) | No – Was <u>not</u> trying to conceive (n=11) |  | 0.58<br>(0.45, 0.66) |
|  | Yes (n=30) | 18 (60.0) | 7 (23.3) | 5 (16.6) | 18 (26.1) |  |
|  | No – Was trying to conceive (n=29) | 2 (6.9) | 27 (93.1) | 0 (0.0) | 27 (39.1) |  |
|  | No – Was <u>not</u> trying to conceive (n=10) | 2 (20.0) | 2 (20.0) | 6 (60.0) | 6 (8.7) |  |
|  | **Tested for a sexually transmitted disease 6 months before pregnancy (n=79)** | Yes (n=5) | No (n=74) |  |  | 0.23<br>(0.02, 0.44) |
|  | Yes (n=21) | 4 (19.0) | 17 (80.9) |  | 4 (5.1) |  |
|  | No (n=58) | 1 (1.7) | 57 (98.2) |  | 57 (72.2) |  |
|  | **Visit a dentist 12 months before pregnancy (n=79)** | Yes (n=29) | No (n=50) |  |  | 0.25<br>(0.04, 0.46) |
|  | Yes (n=36) | 18 (50.0) | 18 (50.0) |  | 18 (22.5) |  |
|  | No (n=43) | 11 (25.5) | 32 (74.4) |  | 32 (40.5) |  |
|  | **Check immunisations 12 months before pregnancy (n=79)** | Yes (n=11) | No (n=68) |  |  | 0.20<br>(-0.01, 0.40) |
|  | Yes (n=28) | 7 (25.0) | 21 (75.0) |  | 7 (8.9) |  |
|  | No (n=51) | 4 (7.8) | 47 (92.1) |  | 47 (59.5) |  |

|  |  |  |  |  |  |
| --- | --- | --- | --- | --- | --- |
|  | **Seeing any health professional about becoming pregnant 12 months before pregnancy (n=54)** | *Yes (n=16)* | *No (n=38)* |  | 0.52<br>(0.31, 0.73) |
|  | *Yes (n=27)* | 15 (55.5) | 12 (44.4) | 15 (27.8) |  |
|  | *No (n=27)* | 1 (3.7) | 26 (96.3) | 26 (48.1) |  |

\* only asked to those participants who reported having ever smoked.

Among couples who had reported ever drinking alcohol, their mean alcohol consumption in the 3 months before pregnancy was dissimilar (data not shown in table). The mean weekday consumption for pregnant females was 1.2 standard drinks per day (SD 1.7) while for non-pregnant partners it was 2.5 (SD 3.5). Within couples this represented very low similarity (ICC: 0.22). Alcohol consumption on weekends was reported by pregnant females as 2.6 standard drinks (SD 3.2) in the three months preconception and non-pregnant partners reported 4.9 standard drinks (SD 5.1), demonstrating poor agreement (ICC: 0.19).

### Participant preconception health information-seeking and information sources

Approximately one quarter (26.3%) of couples reported similarities in partners attempting and succeeding in preconception health information-seeking in the 12 months prior to pregnancy (see Table 3), with 51.2% of pregnant females who sought and found information also having partners reporting the same. A further 43.9% of pregnant females who reported seeking or finding information had partners who did not report doing so. Regarding the sources of preconception health information couples sought and found, almost half of the couples (47.6%) had both partners indicate the information related to themselves and their partner. The highest agreement in preconception health mass information sources accessed by both partners was for the internet (90.5%) and for the sources not accessed by both partners was for magazines (90.5%). Interpersonal information sources with the highest agreement in use among couples were GPs (42.9%) and family and friends (14.3%) while non-use agreement was highest for naturopaths (100.0%), pharmacists (90.5%), and midwives (85.7%). For pregnant females reporting information from their GP, 52.6% had a partner report not receiving information from a GP.

**Table 3:**
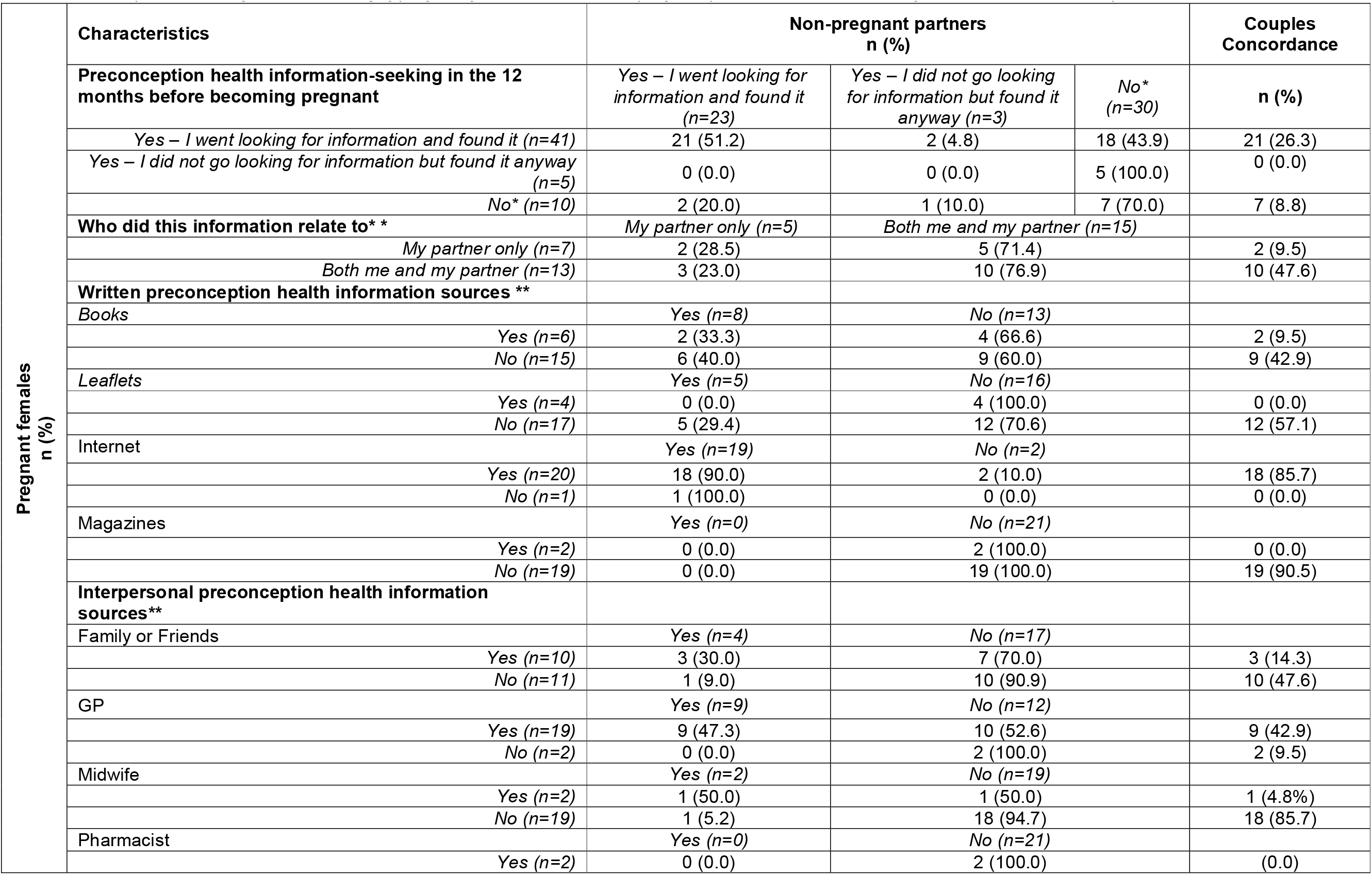
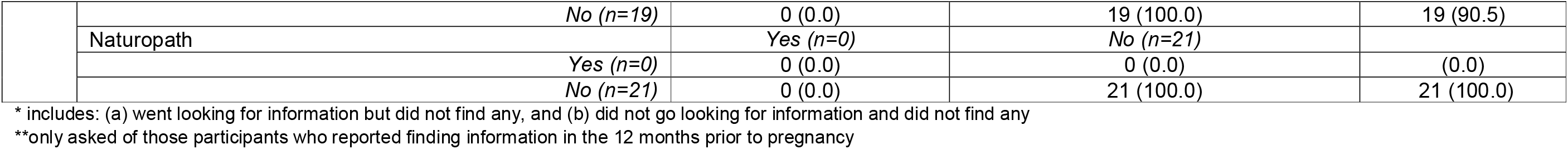
Preconception health information seeking of pregnant females and their non-pregnant partners and concordance of characteristics within couples, N = 80.

### Participant preconception health information discussions

Pregnant females and non-pregnant partners reported the preconception topics discussed with their GPs, other health professionals, or family and friends (see Table 4). Overall, there was a greater concordance in not having discussions with GPs on preconception topics including those explicitly covered within clinical guidelines. The topics with the highest concordance of discussions occurring were for folic acid (10.0%), immunisation (8.7%), and stopping contraception (8.7%). The most common concordance in discussions with GPs not occurring for both partners was for omega 3 fatty acid supplementation (95.0%) and multivitamin use (90.0%). Where pregnant females had discussed a topic with a GP, between 68.1% and 100.0% of their non-pregnant partners had not reported a similar discussion. This disparity was more pronounced for other health professionals, with a maximum 2.5% concordance in reporting a topic for both pregnant females and non-pregnant partners, specifically for discussions about smoking, stopping contraception, and sexually transmitted infections. It was most common for discussions about preconception health topics not to occur between pregnant females or non-pregnant partners and other health professionals or family and friends.

**Table 4:** Preconception health topics discussed with health professionals by pregnant females and their non-pregnant partners and concordance of characteristics within couples, N = 80.

| Preconception health topics | Pregnant Females (n=80) | GP |  |  | Other health care providers* |  |  | Family and friends |  |  |
| --- | --- | --- | --- | --- | --- | --- | --- | --- | --- | --- |
|  |  | Non-pregnant partners (n=80) |  | Couple concordance | Non-pregnant partners (n=80) |  | Couple concordance | Non-pregnant partners (n=80) |  | Couple concordance |
|  |  | Yes | No |  | Yes | No |  | Yes | No |  |
|  |  | n (%) | n (%) | n (%) | n (%) | n (%) | n (%) | n (%) | n (%) | n (%) |
| Eating a healthy diet | Yes | 2 (9.5) | 19 (90.4) | 2 (2.5) | 0 (0.0) | 4 (100.0) | 0 (0.0) | 0 (0.0) | 10 (100.0) | 0 (0.0) |
|  | No | 5 (8.4) | 54 (91.5) | 54 (67.5) | 2 (2.6) | 74 (97.3) | 74 (92.5) | 5 (7.1) | 65 (92.8) | 65 (81.2) |
| Immunisation | Yes | 7 (28.0) | 18 (72.0) | 7 (8.7) | 0 (0.0) | 8 (100.0) | 0 (0.0) | 0 (0.0) | 6 (100.0) | 0 (0.0) |
|  | No | 4 (7.2) | 51 (92.7) | 51 (63.7) | 4 (5.5) | 68 (94.4) | 68 (85.0) | 4 (5.4) | 70 (94.5) | 70 (87.5) |
| Smoking | Yes | 4 (20.0) | 16 (80.0) | 4 (5.0) | 2 (40.0) | 3 (60.0) | 2 (2.5) | 2 (33.3) | 4 (66.6) | 2 (2.5) |
|  | No | 8 (13.3) | 52 (86.6) | 52 (65.0) | 2 (2.6) | 73 (97.3) | 73 (91.2) | 2 (2.7) | 72 (97.3) | 72 (90.0) |
| Alcohol | Yes | 4 (21.0) | 15 (78.9) | 4 (5.0) | 1 (14.3) | 6 (85.7) | 1 (1.2) | 2 (20.0) | 8 (80.0) | 2 (2.5) |
|  | No | 6 (9.8) | 55 (90.1) | 55 (68.7) | 3 (4.1) | 70 (87.5) | 70 (87.5) | 2 (2.8) | 68 (97.1) | 68 (85.0) |
| Physical activity | Yes | 4 (19.0) | 17 (80.9) | 4 (5.0) | 0 (0.0) | 6 (100.0) | 0 (0.0) | 2 (25.0) | 6 (75.0) | 2 (2.5) |
|  | No | 4 (19.0) | 55 (93.2) | 55 (68.7) | 1 (1.3) | 73 (91.2) | 73 (91.2) | 4 (5.5) | 68 (94.4) | 68 (85.0) |
| Stopping contraception | Yes | 7 (31.8) | 15 (68.1) | 7 (8.7) | 2 (50.0) | 2 (50.0) | 2 (2.5) | 0 (0.0) | 3 (100.0) | 0 (0.0) |
|  | No | 1 (1.7) | 57 (98.2) | 57 (71.2) | 0 (0.0) | 0 (0.0) | 0 (0.0) | 1 (1.3) | 76 (98.7) | 76 (95.0) |
| Folic acid | Yes | 8 (26.6) | 22 (73.3) | 8 (10.0) | 0 (0.0) | 11 (100.0) | 0 (0.0) | 1 (7.7) | 12 (92.3) | 1 (1.2) |
|  | No | 0 (0.0) | 50 (100.0) | 50 (62.5) | 0 (0.0) | 69 (100.0) | 69 (86.2) | 1 (1.5) | 66 (98.5) | 66 (82.5) |
| A healthy weight | Yes | 3 (20.0) | 12 (80.0) | 3 (3.7) | 0 (0.0) | 5 (100.0) | 0 (0.0) | 1 (16.6) | 5 (83.3) | 1 (1.2) |
|  | No | 5 (7.7) | 60 (92.3) | 60 (75.0) | 1 (1.3) | 74 (98.6) | 74 (92.5) | 0 (0.0) | 74 (100.0) | 74 (92.5) |
| Sexually transmitted infections | Yes | 5 (26.3) | 14 (73.6) | 5 (6.2) | 2 (40.0) | 3 (60.0) | 2 (2.5) | 0 (0.0) | 1 (100.0) | 0 (0.0) |
|  | No | 3 (4.9) | 58 (95.0) | 58 (72.5) | 1 (1.3) | 74 (98.6) | 74 (92.5) | 2 (2.5) | 77 (97.4) | 77 (96.2) |
| Ordinary multivitamin | Yes | 0 (0.0) | 2 (100.0) | 0 (0.0) | 0 (0.0) | 1 (100.0) | 0 (0.0) | 1 (50.0) | 1 (50.0) | 1 (1.2) |
|  | No | 6 (7.6) | 72 (92.3) | 72 (90.0) | 1 (1.2) | 78 | 78 (97.5) | 0 (0.0) | 78 | 78 (97.5) |
|  |  |  |  |  |  | (98.7) |  |  | (100.0) |  |
| *Omega 3* | *Yes* | 0 (0.0) | 2 (100.0) | 0 (0.0) | 0 (0.0) | 5<br>(100.0) | 0 (0.0) | 0 (0.0) | 2<br>(100.0) | 0 (0.0) |
|  | *No* | 2 (2.5) | 76 (97.4) | 76 (95.0) | 0 (0.0) | 75<br>(100.0) | 75 (93.7) | 1 (1.2) | 77<br>(98.7) | 77 (96.2) |
| *Contraception/fertility* | *Yes* | 5 (18.5) | 22 (81.4) | 5 (6.2) | 6 (54.5) | 5 (45.4) | 6 (7.5) | 0 (0.0) | 2<br>(100.0) | 0 (0.0) |
|  | *No* | 4 (7.5) | 49 (92.4) | 49 (61.2) | 0 (0.0) | 69<br>(100.0) | 69 (86.2) | 3 (3.8) | 75<br>(96.1) | 75 (93.7) |

\*Other HCP includes Midwife, obstetrician/gynaecologist, or naturopath

## Discussion

This study presents an analysis of concordance in pregnancy planning as well as preconception health behaviours and information-seeking among reproductive partners and the findings offer novel insights into the shared preconception experience of couples. Firstly, the couples in this analysis did not report a high degree of concordance in pregnancy planning, as defined by the LMUP. The greatest proportion of discordance was between one partner planning pregnancy and the other reproductive partner indicating ambivalence. This finding was greater than a US study which found 15% of couples with discordant pregnancy intentions [23], although the measure for pregnancy intentions in this previous study was based on questions of perceptions of wantedness and timing only. In contrast, the LMUP defines an individual’s pregnancy intentions as ambivalent when they present mixed feelings towards pregnancy, or their expressed intentions do not completely align with their actions or contraceptive behaviour [18, 20]. The LMUP’s more nuanced approach to defining pregnancy intention may provide new opportunities for exploring pregnancy preparedness within a couple as well as among individuals. However, the implication of the discordance identified through this first analysis of couple’s data using the LMUP requires furthers study, as the ambivalence could be influenced by any of the core items of the LMUP (i.e., contraception use, timing, intention, desire for a baby, partner discussion and preparatory health behaviours). Moreover, this high proportion of ambivalence among at least one reproductive partner in a couple highlights the ongoing challenge for health promotion and clinical communities to enable pregnancy planning and positive preconception health behaviours, to achieve the best possible outcome for both partners and their future child.

Participating couples also demonstrated inconsistent engagement with health services. Research investigating how to better prepare health professionals to deliver preconception care has been growing over recent years [7], but this finding highlights the need to move the approach to strengthening preconception health interventions beyond the clinical encounter. This includes stronger policy actions to address upstream drivers of critical factors impacting preconception health (e.g., housing, exposure to pollution, food security) as well as health promotion campaigns to educate reproductive-aged individuals about modifiable preconception risk factors that can impact pregnancy, foetal and offspring outcomes [1, 2, 8, 24]. Solely focusing on improving preconception care delivered through clinical encounters requires future parents to first present to clinical services, and it is possible that if they do not have immediate health concerns and are unaware of the importance of preconception health, they may not. If this is the case, couples may be unlikely to present together for consultations in which opportunistic preconception health discussions may occur (e.g., contraception medication prescription renewals, chronic condition reviews). As such, in these consultations a clinician can still enquire about partner health and support if pregnancy planning is discussed and encourage a subsequent appointment for individual assessment and care. It may not be necessary for both partners to receive care together, just that they both receive the care they need to prepare for pregnancy. Equally, many modifiable preconception health risk factors do not necessarily require input from a clinician (e.g. healthy eating, healthy body weight, regular exercise, reduced alcohol and nicotine use, etc.) for couples to address, and others may be outside the remit of either health professional or couple to fully impact (e.g., housing, environmental exposures) [1, 2, 8, 24].

The analysis found high rates of internet-based information-seeking among both partners in a couple and that where they found preconception health information, less than half of couples reported both of them finding information related to them and their partner. In Australia, a previous government-supported website providing preconception health information has been defunded and since replaced by another agency [25, 26] while simulated online searches for preconception care services have found numerous private clinics providing preconception health information [27]. The new government-supported website does cover most (but not all) modifiable risk factors and critical topics [28], there is still a greater focus on maternal content in preference to information for male reproductive partners. The reliability and quality of the information sources available on independent clinical websites need further investigation. As only two thirds of services identified for this study refer to male partners among their services [27], it would be unsurprising if this imbalance held true for their public-facing information. Ultimately, government agencies, private clinics and other reputable sources must ensure accurate, current information relevant to both reproductive partners is available to support preconception health behaviour change without clinician involvement while also directing to clinical services where it is needed.

Health professionals were not consistent sources of preconception health information among couples, even when they were accessed by couples who sought preconception health information. GPs were most frequently identified as source of preconception health information, however more than half of pregnant females who reported receiving information from their GP had partners who did not. This aligns with previous research whereby GPs have reported relying on pregnant females to convey preconception health information to their partners [16], but also may reflect a lack of understanding among GPs regarding the importance of paternal preconception exposures [29]. Despite these limitations, GPs still appear to be playing one of the most prominent preconception care roles among health professions. However, there has been substantial research demonstrating willingness among a wide range of health professions (e.g. midwives, maternal child health nurses, pharmacists, naturopaths [7]) to deliver preconception care and support preconception health behaviour change. There is also unrealised value of approaching preconception care services through the lens of preventive health, whereby every contact with the health system counts [8]. However, further research is needed to fully understand the scope of preconception care those professions can deliver, their readiness to provide care to both partners, and couples’ perceptions on the appropriateness of each profession to deliver preconception care.

An important overall difference in preconception behaviour and pregnancy planning is the lower engagement from non-pregnant partners. This was most pronounced with the overall figure of 1 in 5 pregnant females with planned pregnancy whose partners were ambivalent. But there were also many behaviours reported by pregnant females but not reported by their non-pregnant partners, despite evidence of the behaviour contributing risk to pregnancy and offspring outcomes irrespective of the partner. One such example is alcohol consumption [1, 2]. Maternal preconception exposure to alcohol is associated with lower conception rates, an increased risk of miscarriage, reduced embryonic growth, and increased risk of neural tube defects and other congenital anomalies [2]. Even pregnant females consuming as little as two alcoholic drinks per day during the preconception period, as was seen in this study’s sample, increases risk for small for gestational age infants [2]. While there has been less investigation into the impacts of paternal preconception alcohol exposure, current evidence suggests eight or more standard drinks per week may impact androgen-dependent reproductive development among male offspring [1]. Based on current evidence, there are positive effects associated with both partners limiting their alcohol intake prior to pregnancy. Yet, our study indicates not only a higher intake of alcohol among some non-pregnant partners but also less discussion about alcohol intake with those partners compared to the pregnant females in the same couple. A similar pattern can be seen for healthy eating in which the diet quality and nutrient intake of pregnant females can positively impact fecundity, miscarriage risk, birth weight, and size for gestational age [2]. Likewise, paternal healthy eating can improve fertility rates and premature birth risk [1]. Even paternal diet as an adolescent can impact offspring growth patterns [1]. Yet there was an inconsistent prevalence of preconception health information about health eating given to pregnant females and their non-pregnant partners. While this pattern may in part be explained by poor health service engagement among adult males [30], leaving this information and behaviour gap unaddressed only places greater burden on the pregnant female to source and share relevant preconception health information and support preconception behaviour change in their non-pregnant partner. Such an approach only reinforces the pervasive gender imbalance occurring in preconception care [31]. It also overlooks the emerging evidence that preconception health behaviour change for both partners in a couple planning pregnancy is influenced by the perceived beliefs of their counterpart, more so than by those of a health professional [13, 14].

## Strengths and Limitations

The results must be considered within the context of the study’s limitations. The sample size restricted application of concordance tests like Cohen’s kappa for some variables due to lower response numbers for those items causing sparse categories and imbalanced distributions. These limitations can cause the expected agreement to fluctuate, making the use of Cohen’s kappa somewhat unstable and sensitive to small changes in the data. It is also important to note that the LMUP used in this study included five of the six nominated questions due to an error in the online survey logic. The scoring for this instrument was adjusted accordingly, but this change may have affected the reliability of the score. Overall, despite these limitations, this is the first analysis of reproductive couple dyads that investigates the concordance between their preconception health intentions, behaviours and information-seeking.

## Conclusion

This study highlights significant discordance in pregnancy intention, preconception health behaviours, and preconception health information-seeking within couples who become pregnant, with lower levels of all three areas noted for the non-pregnant partners. This difference supports the growing calls for better engagement with males about their direct and indirect role and influence in healthy pregnancies and healthy children. Importantly, this study also highlights the need to develop new approaches to preconception health interventions that include both reproductive partners and ensure pregnancy preparation is, where appropriate, shared equally within a couple. The impact of these findings warrant further study, and should be carefully considered by clinicians, health service managers and policymakers.

## Data availability

Ethical clearance did not include public sharing of data. A de-identified dataset is available from the authors upon reasonable request and subject to approval from the overseeing ethics committee.

## Acknowledgements

The authors acknowledge the contributions of Dr Jennifer Hall and Dr Erica McIntyre in drafting the survey instrument used in this study.

